# Content-based image retrieval assists radiologists in diagnosing eye and orbital mass lesions in MRI

**DOI:** 10.1101/2024.07.24.24310920

**Authors:** Josef Lorenz Rumberger, Winna Lim, Benjamin Wildfeuer, Elisa Birgit Sodemann, Augustin Lecler, Simon Stemplinger, Ahi Sema Issever, Ali Sepahdari, Sönke Langner, Dagmar Kainmueller, Bernd Hamm, Katharina Erb-Eigner

## Abstract

Diagnosing eye and orbit pathologies through radiological imaging presents considerable challenges due to their low prevalence, the extensive range of possible conditions, and their variable presentations, necessitating substantial domain-specific expertise. This study evaluates whether a ML-based content-based image retrieval (CBIR) tool, combined with a curated database of orbital MRI cases with verified diagnoses, can enhance diagnostic accuracy and reduce reading time for radiologists diagnosing eye and orbital pathologies. It explores whether this tool alone, or in combination with status quo reference tools (e.g. Radiopaedia.org, StatDx) provides these benefits. In a multi-reader, multi-case study involving 36 radiologists and 48 retrospective orbital MRI cases, participants diagnosed eight cases: four using status quo reference tools and four with the addition of the CBIR tool. Analysis using linear mixed-effects models revealed significant improvements in diagnostic accuracy when using the CBIR tool alone (55.88% vs. 70.59%, p = 0.03, odds ratio = 2.07) and an even greater improvement when used alongside status quo tools (55.88% vs. 83.33%, p = 0.02, odds ratio = 3.65). Reading time decreased when using the CBIR tool alone (334s vs. 236s, p < 0.001) but increased when used in conjunction with status quo tools (334s vs. 396s, p < 0.001). These findings indicate that CBIR tools can significantly enhance diagnostic accuracy for eye and orbit diagnostics, though their impact on reading time varies.

## Introduction

Inaccurate diagnoses in medical imaging reports are a burden to the patient and the healthcare system^1^. Reading MRI scans of patients with eye and orbit diseases poses a particular diagnostic challenge due to the rarity of these lesions. Most radiologists lack profound experience reading these cases or they may find it difficult to recall imaging features from past cases. Radiologists specialized in the eye and orbit area are also rare, thus these cases are often read by general radiologists or neuroradiologists, increasing the probability for diagnostic inaccuracies. Additionally, the high number of distinctive tissue types in the orbit enables a variety of orbital pathologies, increasing the number of possible differential diagnoses to consider.

Although large, multi-center studies describing the diagnostic accuracy of eye and orbital lesions are lacking, it has been reported for lacrimal gland lesions that the degree of correspondence between image-based diagnosis and histopathologic diagnosis is only moderate (Cohen’s kappa=0.451, p <0.001)^2^. Other studies found that diagnostic errors occur at an average rate of 3-4%, with a 32% retrospective error rate for interpretation of abnormal studies^3^. These challenges may delay diagnosis and treatment or expose patients to potentially unnecessary biopsies and treatments, which can cause harm and be costly^1^.

Content-based image retrieval (CBIR) methods retrieve similar images from a database based on a query image, by comparing visual features like color, texture, and shape, rather than metadata or text^4^. In the context of Radiology, CBIR systems allow radiologists to retrieve relevant cases from a curated database with clinical or histopathological validation, based on visual similarity with supplied patient query images. Given the cases and their associated diagnoses retrieved by the CBIR system, radiologists may be able to give better informed and more accurate diagnoses. Previous studies on CBIR showed increases in diagnostic accuracy, particularly for diagnosing interstitial lung diseases on CT scans^5–8^. However, these studies often did not compare CBIR with status quo reference tools (e.g. StatDx, radiopaedia.org, etc.)^7,8^, and involved a small number of participants, albeit many cases per participants. Notably absent is research on CBIR’s effectiveness in challenging MRI diagnoses and other organ systems where retrieval of reference cases can be crucial and time consuming.

Thus, our study seeks to close this gap by evaluating whether a CBIR system can improve diagnostic accuracy and reading time for diagnosing challenging eye and orbital pathologies. We developed an ML-based CBIR tool and conducted a retrospective study involving 36 radiologists and 48 orbital MRI cases to assess its effectiveness across a wide range of experience levels and orbital pathologies.

## Methods

### Ethics statement

This retrospective study was approved by the institutional review board of Charité University Medicine under ethics application code EA121422. The study was conducted in strict accordance with relevant guidelines and regulations. Written consent was obtained from radiologists participating in the study, informed consent from patients was waived due to the retrospective character of the study. All data were completely anonymized before inclusion.

### Orbital pathologies datasets

For developing the CBIR machine learning (ML) model and the database, we collected anonymized data from patients with eye and orbit pathologies who were diagnosed between 2012 and 2022 at Charité University Medicine, Hôpital Fondation Rothschild, and Scripps Hospital La Jolla (**Fig. 1 a**). The inclusion criteria required clinical or histopathological confirmation of the diagnosis verified through multidisciplinary clinical assessments, visible lesions on the respective MRI scans, scans performed prior to any therapeutic treatment, and sufficient image quality. For the ML model development, 3D regions of interest (ROIs) were annotated as bounding boxes around each lesion by three expert radiologists in a consecutive non-blinded manner. For annotation and review of the dataset we build tailormade data annotation software (based on Ruby-on-rails and the OHIF DICOM viewer). The following routinely acquired MRI sequences were annotated: T1-weighted spin echo sequences before and after intravenous contrast agent administration, T2-weighted sequences with and without fat suppression, and Fluid-Attenuated Inversion Recovery sequences. Sequences were acquired with a range of different scanners: Siemens (Skyra, Aera, Avanto, Magnetom Amira, Vida), Philips (Ingenia, Intera, Symphony), Toshiba (Titan) and GE (Optima, Signa). Field strength varied between 1.5T and 3T depending on the scanner. Data from Charité University Medicine and Hôpital Fondation Rothschild was split into training and validation cases, with the validation dataset being constructed by taking 10% of cases of each pathology to ensure a representative sample. The Scripps dataset was used as an external test dataset.

**Fig. 1.**
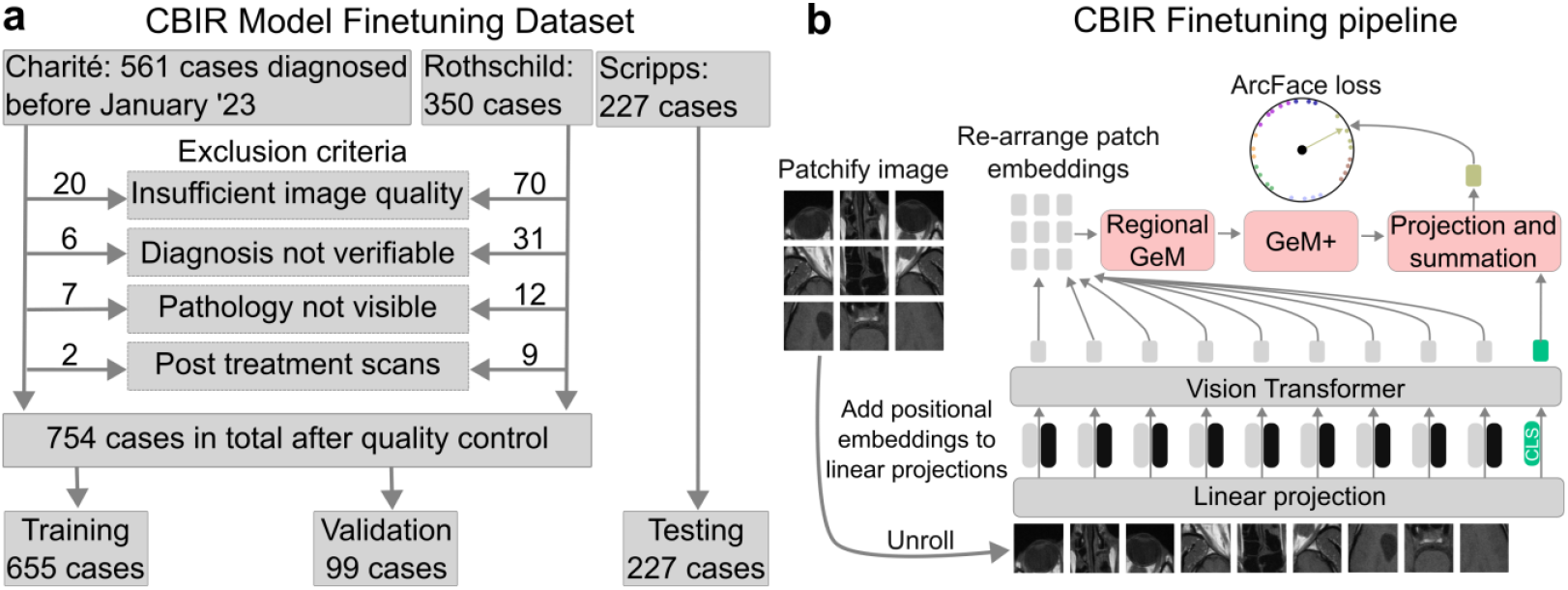
Dataset composition and finetuning. **a**, for the CBIR model we gathered data from 3 sources and excluded cases based on a range of quality control measures. **b**, we used the training dataset to finetune a vision transformer with class token (CLS), Regional Generalized Mean (GeM) Pooling and GeM pooling with manual hyperparameter tuning (GeM+) for image retrieval, by optimizing the ArcFace loss.

For the reader study, data with similar characteristics, but diagnosed after January 2023 were collected at Charité University Medicine. The dataset included 142 cases, spanning 28 pathologies which were a subset of the pathologies present in the training dataset. Six sets of eight cases were randomly sampled for the reader study, such that each set consisted of cases with eight distinct pathologies without repetition (**Fig. 2. a-b**). This sampling procedure resulted in 48 cases spanning 20 different pathologies, with the pathology distribution shown in **Fig. 2. c**. The 48 sampled patients had an average age of 43±24 years and 48% were female.

**Fig. 2.**
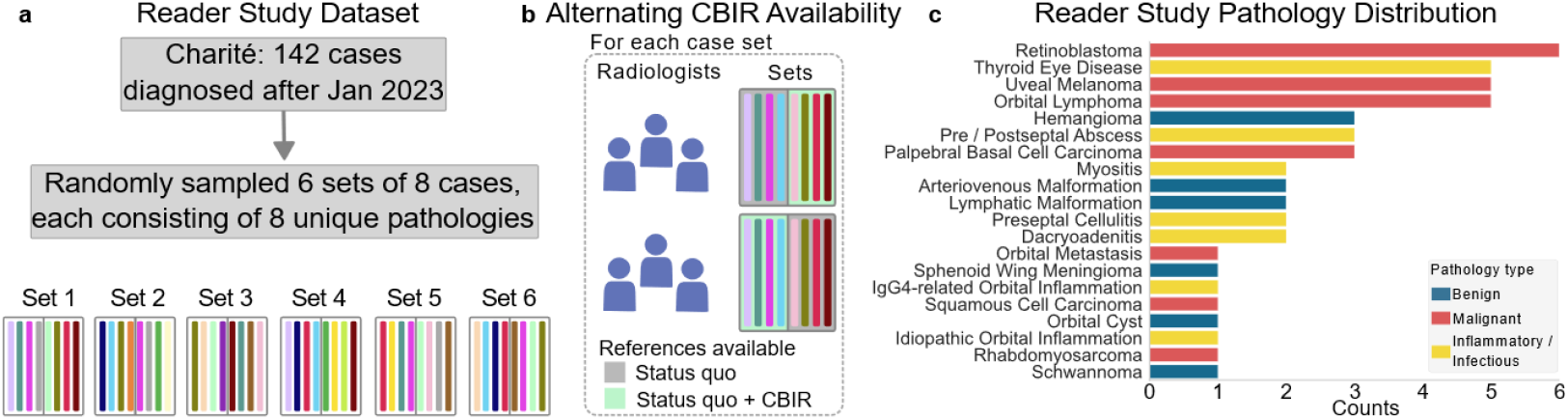
Reader study dataset composition. **a**, for the reader study we only used cases from Charité University Medicine, diagnosed after the cases in the finetuning dataset. **b**, cases were sampled such that each case set contained 8 distinct diagnoses, and the sets were read by 6 radiologists with alternating CBIR availability. **c**, the randomly sampled 48 cases span 20 distinct diagnoses of different types.

### Content-Based Image Retrieval Tool

The CBIR tool is seamlessly integrated into the PACS viewer and accessible to eligible radiologists with one click on a dedicated button in the PACS (**Fig. 3 a**). To use the CBIR tool, users navigate to a sequence slice where the pathology is clearly visible, then click on the button which opens the web application that shows a range of pathologies, sorted by image similarity (**Fig. 3 b**). The user interface enables exploration of several cases across 77 verified eye and orbit pathologies in seven anatomical subregions (preseptal space, globe, optic nerve, intraconal, extra ocular muscles, extraconal, lacrimal gland, subperiosteal space and bony orbit). The CBIR algorithm employs an ML model that compares the uploaded radiology sequence slice with those in the database, ranking them by similarity. The algorithm is based on the DinoV2 self-supervised learning framework^9,10^, whose pre-trained checkpoint was further trained on publicly available radiology datasets^11^ (see **Suppl. S3** for details). A head comprising Regional Generalized Mean (Regional GeM) Pooling and GeM+ Pooling^12^ was added to extract features from the patch embeddings, which were then combined with CLS token features into a single embedding vector. The model was finetuned on the ArcFace image-retrieval objective^13^ using the CBIR fine-tuning dataset (**Fig. 1 b**). More details on the data pre-processing steps and the model performance are presented in **Suppl. S2-S4 and Suppl. Fig. 1**. The ML model was developed using PyTorch (version 2.3.0) and Python (version 3.10).

**Fig. 3.**
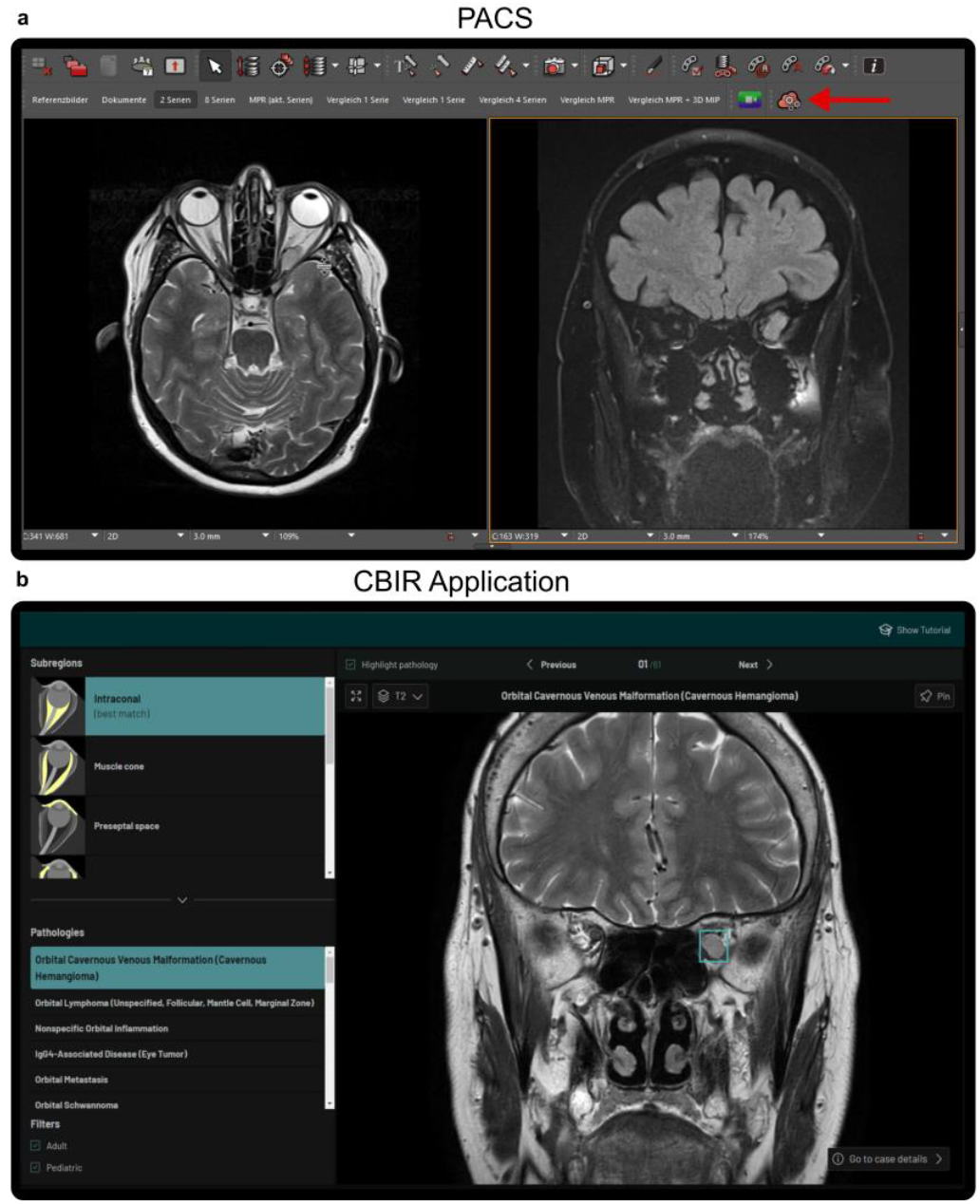
PACS integrated CBIR application. **a**, the PACS viewer environment, with the button starting the CBIR tool highlighted with a red arrow. **b**, the CBIR web application with the search results for the slice shown on the right in **a**. The pathology is highlighted with a cyan box in **b**.

### Study population

The study was conducted in March and April 2024 at Charité University Medicine. Eligible for the study were radiologists with experience in reading MRI exams. 36 radiologists were randomly recruited for the study, who covered a representative cross section of the department (**Tab. 1 a**), working in a range of medical roles (**Tab. 1 b**), having varying job tenure (**Tab.1 c**). Prior experience in reading orbital MRI cases was low (**Tab. 1 d**), with 28 of 36 participants having either no or only little prior experience.

**Tab. 1.** Study participant demographics. Relative number of participants stratified by sex (**a**), medical role (**b**), tenure (**c**) and prior experience in reading orbital MRIs (**d**). Fractions of total number of participants in parentheses.

| Demographic | Share of participants |
| --- | --- |
| **a \| Sex** |  |
| Female | 41.67 (15/36) |
| **b \| Medical Role** |  |
| Resident | 44.44 (16/36) |
| Board-certified | 27.78 (10/36) |
| Senior | 27.78 (10/36) |
| **c \| Tenure** |  |
| 0 - 5 years | 44.44 (16/36) |
| 6 - 10 years | 27.78 (10/36) |
| 11 - 15 years | 11.11 (4/36) |
| >15 years | 16.67 (6/36) |
| **d \| Prior exp. in orbital MRI** |  |
| No exp. | 19.44 (7/36) |
| Little exp. | 58.33 (21/36) |
| Sufficient exp. | 22.22 (8/36) |

### Reader evaluation

In total 36 participants each diagnosed a set of eight cases only based on the MRI scans (**Fig. 2 a**), four with and four without the CBIR tool available. Other status quo reference tools like radiopaedia.org, StatDx or Google were available throughout the study. Half of the participants had the CBIR tool available for the first four cases, whereas the other half for the last four cases. Each individual case was read by six randomly selected participants with alternating availability of the CBIR tool (**Fig. 2 b**). Before the participants read cases with the CBIR tool, they went through a short tutorial and were allowed to test the tool by diagnosing a case with a pathology not present in the reader study dataset. In addition, they were allowed to ask questions of the experimenter regarding the CBIR tool. Cases were read on radiology workstations within a standard PACS environment. After each case, the participants were asked to give their diagnosis in free-text form, rate the perceived difficulty, provide their confidence level in the diagnosis, and the reference tools that they used.

Confidence levels and difficulty ratings were assessed using a 4-point Likert scale, designed as a forced-choice format without a neutral option to encourage definitive responses. For confidence, participants responded to the statement ‘You are confident that your diagnosis is correct.’ with one of the following options: ‘Strongly agree,’ ‘Somewhat agree,’ ‘Somewhat disagree,’ or ‘Strongly disagree.’ For difficulty ratings, they answered the question ‘How would you assess the difficulty level of this case?’ with one of the following choices: ‘Very difficult,’ ‘Difficult,’ ‘Easy,’ or ‘Very easy.’ A person instructing the participants and taking time measurements was in the room during the session. After the measurements were completed, an eye and orbit radiology specialist with over 15 years of expertise with access to additional clinical information on each case, assessed the diagnoses given by the participants in a fully blinded manner. The evaluation was based on the criterion that the diagnosis was sufficiently correct to ensure the accurate administration of downstream treatment, meaning only clinically significant errors were counted as being incorrect (more details in **Suppl. S1**). This assessment considers that the classification of orbital lesions can vary among centers and countries, thus diagnostic accuracy should not be judged merely on technical correctness, but on its clinical impact on patient management and outcomes.

### Statistical Analysis

Prior to commencement of the study, a power analysis was conducted to determine the number of participants required to detect significant effects (defined as p<0.05) for the endpoints. We reviewed effect sizes from comparable studies and calculated that a sample size of 36 participants and 48 cases, resulting in 288 measurements in total, would allow us to detect effects down to an effect size of Cohen’s D 0.6 at 80% statistical power (more details in **Suppl. S7 and Suppl. Fig. 2**)^14^.

Instead of analyzing if the availability of the CBIR tool had an effect on accuracy and reading times, we focused on the actual reference tools that the participants used for each case, which we measured during the study for each participant and case individually. Therefore, we split reference tool usage into four categories: no reference used, only status quo (only SQ) used, only CBIR used, or both status quo and CBIR used (SQ+CBIR). However, the ‘no reference used’ category was not further analyzed in direct comparison to cases where reference tools were used, as participants refrained from using references only when they immediately and confidently recognized the diagnosis. This introduces a strong selection effect, rendering direct comparisons with tool-assisted observations inappropriate (see **Suppl. S6** and **Suppl. Tab. 3** for details). We analyzed the effect of the CBIR tool on diagnostic accuracy using a logistic mixed effects model, treating individual participants and cases as random effects, and including reference usage, medical roles, tenure, and interaction terms as fixed effects. For analyzing the effect of the CBIR tool on reading times, we employed a linear mixed effects model with the same random and fixed effects. Reading times were log-transformed, to meet the distributional assumption of the model. We excluded fixed effects via a backwards elimination process based on the Akaike information criterion^15,16^. The residuals of the mixed effects models were examined to check if all assumptions were met in accordance with the approach published by Singer et al.^17^. Reported P values are based on two-sided *Student’s t* tests for generalized mixed effects models. Statistical analysis and data visualization were performed using R (version 4.3.3) and Python (version 3.10).

## Results

Participants spent on average (± standard deviation) 01h:03m:57s ± 35m:31s in total on the tutorial, reading the cases and providing the measurements. When not accounting for the reference tools that the participants actually used but only for the ones that were available in the respective study phase, reading times stayed approximately constant (no CBIR 260s, CBIR 257s p=0.09, N=288), while during the CBIR phase participants used reference tools more often (no CBIR 70.14%, CBIR 92.36%) and had a significantly higher diagnostic accuracy (no CBIR 63.19%, CBIR 73.61% p=0.049, N=288) (**Tab. 2 a)**. In addition, having the CBIR tool available slightly increased confidence in the diagnoses (**Tab. 2 b**). No trend is visible on the perceived difficulty of the cases over the study phases (**Tab. 2 c**). Without the CBIR tool available, most participants used radiopaedia.org and Google for finding reference cases, whereas with the CBIR tool available, participants used considerably fewer other reference resources (**Tab. 2 d**). Participants often used only the CBIR tool when it was available and only used additional status quo reference tools in 20.83% of the cases (**Tab. 2 e**). In the following sections, the impact on the diagnostic accuracy and reading times of using only status quo (only SQ) reference tools, only the CBIR reference tool (only CBIR), and using both in conjunction (SQ+CBIR) are analyzed.

**Tab. 2.** Summary statistics split by treatment phase. Unless otherwise stated, data is presented as percentages relative to the total number of measurements. Fractions of total number of measurements in parenthesis. Participants were allowed to use multiple reference tools, so the relative numbers in **d** add up to more than 100%.

| Characteristics | No CBIR | CBIR |
| --- | --- | --- |
| **a \| General** |  |  |
| Reading time [s] | 260±228 | 257±193 |
| Reading time with reference tool [s] | 336±230 | 272±193 |
| Reference tool use | 70.14 (101/144) | 92.36 (133/144) |
| Accurate diagnoses | 63.19 (91/144) | 73.61 (106/144) |
| **b \| Confidence** |  |  |
| Really low confidence | 9.03 (13/144) | 4.17 (6/144) |
| Low confidence | 17.36 (25/144) | 18.06 (26/144) |
| Sufficient confidence | 61.81 (89/144) | 63.19 (91/144) |
| High confidence | 11.81 (17/144) | 14.58 (21/144) |
| **c \| Difficulty** |  |  |
| Really easy | 0.00 (0/144) | 0.00 (0/144) |
| Easy | 37.50 (54/144) | 35.42 (51/144) |
| Hard | 47.22 (68/144) | 48.61 (70/144) |
| Really hard | 14.58 (21/144) | 12.50 (18/144) |
| Not stated | 0.07 (1/144) | 3.47 (5/144) |
| **d \| Reference tools used by participants** |  |  |
| CBIR tool | 0.00 (0/144) | 91.67 (132/144) |
| Radiopaedia | 59.72 (86/144) | 18.06 (26/144) |
| Google | 38.19 (55/144) | 10.42 (15/144) |
| StatDx | 9.03 (13/144) | 1.39 (2/144) |
| Pubmed | 6.94 (10/144) | 0.00 (0/144) |
| Others | 2.08 (3/144) | 0.69 (1/144) |
| **e \| Reference categories** |  |  |
| No reference used | 29.86 (43/144) | 7.64 (11/144) |
| Only SQ | 70.14 (101/144) | 0.69 (1/144) |
| Only CBIR | 0.00 (0/144) | 70.83 (102/144) |
| SQ+CBIR | 0.00 (0/144) | 20.83 (30/144) |

### Impact of CBIR Usage on Diagnostic Accuracy

Diagnostic accuracy significantly improved overall from 55.88% with status quo reference tools only, to 70.59% when using the CBIR tool only (odds ratio=2.07, p=0.03) and to 83.33% when using the CBIR tool in conjunction with status quo tools (odds ratio=3.65, p=0.02, **Suppl. Tab. 1**), which constitutes a 26.32% and a 49.12% relative improvement over the status quo (**Tab. 3 f**).

**Tab. 3.** Diagnostic Accuracy with/out CBIR. Statistics of diagnostic accuracy in percent are shown for measurements where only status quo reference tools were used (Only SQ), where only the CBIR tool was used (Only CBIR) and where both were used (SQ+CBIR). Total numbers as fractions in parentheses. P values indicate significant differences to reference level ‘Only status quo’ and were calculated using logistic mixed effects models with individual readers and patients as random effects. All models were estimated with the full dataset, consisting of 288 measurements in total.

| Characteristics | Only SQ | Only CBIR | P value | SQ+CBIR | P value |
| --- | --- | --- | --- | --- | --- |
| **a \| Difficulty** |  |  |  |  |  |
| Easy | 65.52 (19/29) | 91.18 (31/34) | 0.02* | 100.00 (9/9) | 0.99 |
| Hard | 56.60 (30/53) | 62.00 (31/50) | 0.47 | 75.00 (12/16) | 0.23 |
| Really hard | 40.00 (8/20) | 46.15 (6/13) | 0.86 | 80.00 (4/5) | 0.13 |
| **b \| Pathology Type** |  |  |  |  |  |
| Infl. & Infect. | 55.56 (20/36) | 77.78 (28/36) | 0.055 | 81.82 (9/11) | 0.11 |
| Benign | 43.48 (10/23) | 44.44 (8/18) | 0.93 | 83.33 (5/6) | 0.12 |
| Malignant | 62.79 (27/43) | 75.00 (36/48) | 0.22 | 84.62 (11/13) | 0.23 |
| **c \| Medical Role** |  |  |  |  |  |
| Resident | 62.26 (33/53) | 79.49 (31/39) | 0.09 | 80.95 (17/21) | 0.11 |
| Board-certified | 59.09 (13/22) | 53.13 (17/32) | 0.62 | 75.00 (3/4) | 0.62 |
| Senior | 40.74 (11/27) | 77.42 (24/31) | 0.01* | 100.00 (5/5) | 0.99 |
| **d \| Prior Experience** |  |  |  |  |  |
| No exp. | 52.00 (13/25) | 77.27 (17/22) | 0.10 | 100.00 (6/6) | 0.11 |
| Little exp. | 57.38 (35/61) | 69.81 (37/53) | 0.15 | 79.17 (19/24) | 0.058 |
| Sufficient exp. | 56.25 (9/16) | 66.67 (18/27) | 0.79 | - |  |
| **e \| Tenure** |  |  |  |  |  |
| 0-5 years | 62.26 (33/53) | 79.49 (31/39) | 0.10 | 80.95 (17/21) | 0.11 |
| 6-10 years | 41.67 (10/24) | 61.76 (21/34) | 0.15 | 100.00 (4/4) | 0.90 |
| 11-15 years | 55.56 (5/9) | 50.00 (6/12) | 0.64 | - |  |
| >15 years | 56.25 (9/16) | 82.35 (14/17) | 0.16 | 80.00 (4/5) | 0.44 |
| **f \| Overall** |  |  |  |  |  |
| All | 55.88 (57/102) | 70.59 (72/102) | 0.03* | 83.33 (25/30) | 0.02* |

At the case level, accuracy increased on average with CBIR usage in 21 cases, stayed constant for 18 cases and decreased for 9 cases (**Fig. 4 b** cases above, on and below the isoline). For three cases, diagnostic accuracy declined considerably with the CBIR tool available (from 66.66% without CBIR to 0% with CBIR), which we discuss in more detail in **Suppl. S5**. Accuracy declined with increased perceived difficulty independent of reference tool use, but using the CBIR tool retained a higher accuracy across increasing difficulty levels (**Fig. 4 a, Tab. 3 a, Suppl. Fig. 3 a-b**). Most cases within a pathology were consistently rated with similar difficulty ratings by study participants across experience levels (**Suppl. Fig. 3 b-c**), with the highest difficulty ratings given to arteriovenous malformations, orbital cysts, metastases and schwannomas. Difficulty ratings were relatively consistent across study participants of different prior experience levels (**Suppl. Fig**. **3 b**). We found an increase in diagnostic accuracy from 65.52% with status quo tools only, to 91.18% with the CBIR tool only, a 39% relative increase (p=0.02) for ‘easy’ cases. For ‘hard’ and ‘really hard’ cases, we found similar positive trends (**Tab 3 b**). Stratified by pathology type, the highest increase in accuracy was observed for inflammatory and infectious diseases (only SQ 55.56%, only CBIR 77.78% p=0.055, SQ+CBIR 81.82% p=0.11), albeit not significant.

**Fig. 4.**
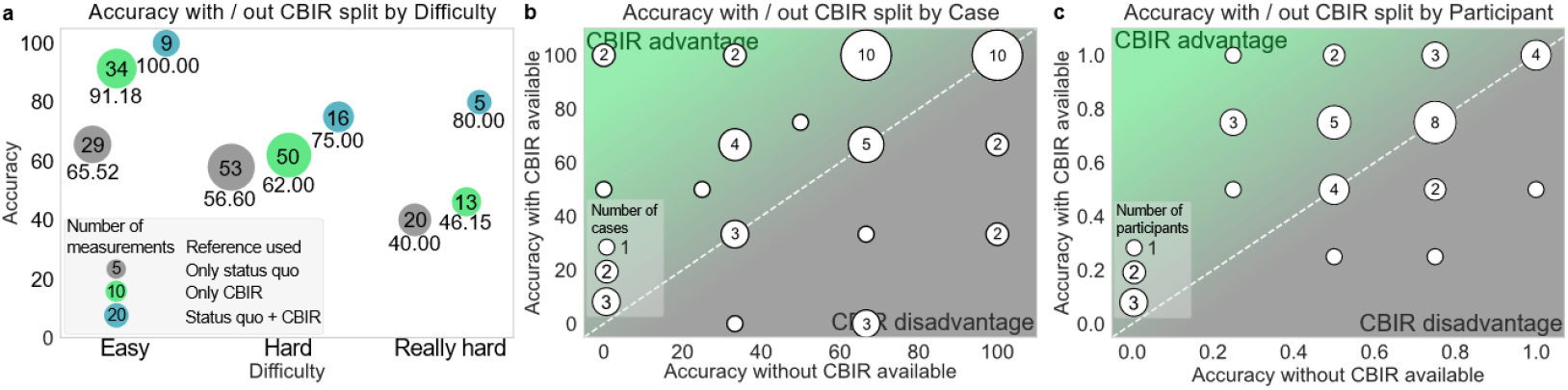
Diagnostic Accuracy. **a**, diagnostic accuracy averaged over individual cases that readers perceived as easy, hard or really hard. **b-c**, diagnostic accuracy with CBIR available (Y-axis) and without CBIR available (X-axis) averaged over individual cases (**b**) and over individual study participants (**c**). Dots above the white isoline indicate higher accuracy with the CBIR tool than without and vice versa. Dot-size indicates the number of measurements (**a**), of cases (**b**) and participants (**c**).

At the participant level, diagnostic accuracy increased on average for 15 study participants, stayed constant for 16 and decreased for 5 (cf. **Fig. 4 c**, participants above, on and below the isoline). Accuracy of participating senior radiologists improved with the CBIR tool (only SQ 40.74%, only CBIR 77.42% p=0.01), whereas accuracy of resident and board-certified radiologists showed positive but insignificant trends (**Tab. 3 c**). Diagnostic accuracy improved the most for participants with no experience (only SQ 52%, only CBIR 77.27% p=0.10, SQ+CBIR 100% p=0.11) and those with little experience (only SQ 57.38%, only CBIR 69.81% p=0.15, SQ+CBIR 79.17% p=0.058), albeit not significantly (**Tab. 3 d**). Accuracy showed a positive trend for all tenure levels, except for the 11-15 years tenure level where it showed a slightly decreasing trend (only SQ 55.56%, only CBIR 50% p=0.64, **Tab. 3 e**).

### Impact of CBIR Usage on Reading Time

Reading time decreased by 29% when using only the CBIR tool compared to only status quo tools (only SQ 334s, only CBIR 236s p<0.001). In contrast, reading time increased by 19% when using CBIR in conjunction with status quo tools (only SQ 334s, SQ+CBIR 396s p<0.001, **Tab. 4 f** and **Suppl. Tab. 2**).

**Tab. 4.** Reading time with/out CBIR. Statistics of reading time averages ± standard deviations in seconds are shown for measurements where only status quo reference tools were used (Only SQ), where only the CBIR tool was used (Only CBIR) and where both were used (SQ+CBIR). P values indicate significant differences to reference level ‘Only status quo’ and were calculated using linear mixed effects models with individual readers and patients as random effects. All models were estimated with the full dataset, consisting of 288 measurements in total.

| Characteristics | Only SQ | Only CBIR | P value | SQ+CBIR | P value |
| --- | --- | --- | --- | --- | --- |
| **a \| Difficulty** |  |  |  |  |  |
| Easy | 202±113 | 158±78 | 0.14 | 260±173 | 0.01* |
| Hard | 357±206 | 271±198 | 0.002* | 462±212 | 0.03* |
| Really hard | 464±317 | 364±144 | 0.41 | 428±215 | 0.94 |
| **b \| Pathology Type** |  |  |  |  |  |
| Infl. & Infect. | 338±201 | 226±112 | 0.005* | 389±242 | 0.10 |
| Benign | 363±240 | 335±304 | 0.40 | 476±278 | 0.34 |
| Malignant | 314±250 | 207±126 | <0.001* | 365±163 | 0.045* |
| **c \| Medical Role** |  |  |  |  |  |
| Resident | 417±278 | 276±232 | <0.001* | 441±223 | 0.046* |
| Board-certified | 208±96 | 228±136 | 0.34 | 205±64 | 0.79 |
| Senior | 273±112 | 195±90 | 0.08 | 360±188 | 0.58 |
| **d \| Prior Experience** |  |  |  |  |  |
| No exp. | 313±160 | 288±181 | 0.75 | 393±140 | 0.19 |
| Little exp. | 377±263 | 236±186 | <0.001* | 397±232 | 0.054 |
| Sufficient exp. | 204±113 | 194±122 | 0.50 | - |  |
| **e \| Tenure** |  |  |  |  |  |
| 0-5 years | 417±278 | 276±232 | <0.001* | 441±223 | 0.049* |
| 6-10 years | 237±104 | 210±126 | 0.58 | 408±179 | 0.29 |
| 11-15 years | 187±89 | 250±133 | 0.32 | - |  |
| >15 years | 286±114 | 188±74 | 0.17 | 198±57 | 0.73 |
| **f \| Overall** |  |  |  |  |  |
| All | 334±230 | 236±172 | <0.001* | 396±215 | <0.001* |

At the case level, reading time decreased when using only the CBIR tool and increased when using it together with SQ tools, for hard cases (only SQ 357s, only CBIR 271s p=0.002, SQ+CBIR 462s p=0.03, **Fig. 5 a, Tab. 4 a**). In addition, we found evidence for a similar effect for malignant lesions (only SQ 314s, only CBIR 207s p<0.001, SQ+CBIR 365s p=0.045) and a decrease in reading times for inflammatory and infectious lesions when using only the CBIR tool (only SQ 338s, only CBIR 226s p=0.005, **Fig. 5 b, Tab. 4 b**).

**Fig. 5.**
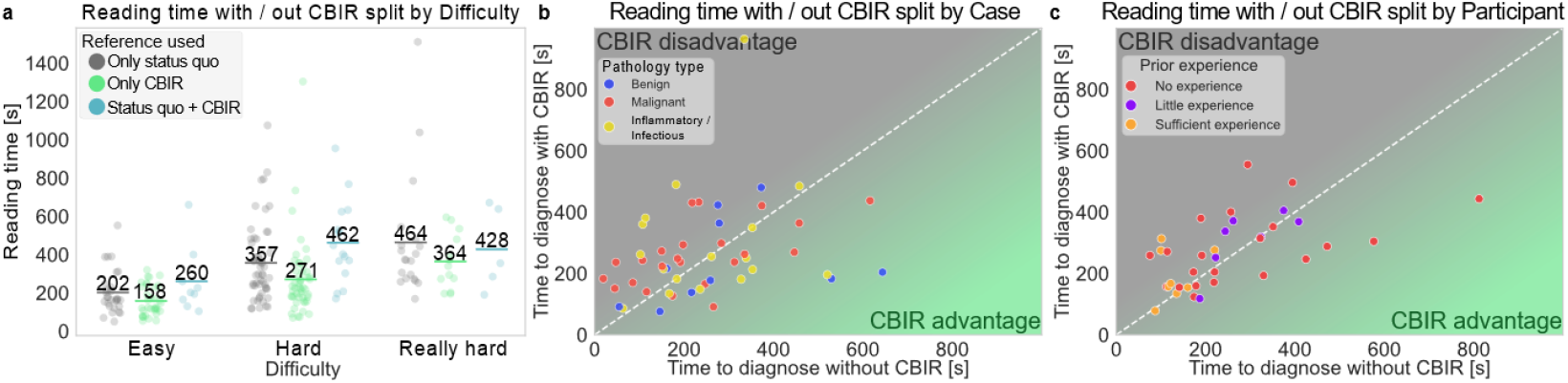
Reading time. **a**, reading time split by perceived difficulty and use of the CBIR tool with averages overlayed. **b-c**, reading time with CBIR available (Y-axis) and without CBIR available (X-axis) split by cases (**b**) and study participants (**c**). Dots below the white isoline indicate a lower reading time with the CBIR tool than without and vice versa, dots on the isoline.

At the participant level, resident radiologists benefited the most from the CBIR tool (only SQ 417s, only CBIR 276s p<0.001, **Tab. 4 c**). In addition, the decrease in reading time was the strongest for participants with little experience (only SQ 377s, only CBIR 236s p<0.001, **Fig. 5 c, Tab. 4 d**). Reading times among participants of different tenure levels decreased the most for the 0-5 years of tenure group, with a relative decrease of 31% (only SQ 417s, only CBIR 276s p<0.001), while they showed an increase when both CBIR and SQ tools were used together (only SQ 417s, SQ+CBIR 444s p=0.049, **Tab. 4 e**).

## Discussion

Our results indicate a significant positive impact on diagnostic accuracy with high effect sizes when using the CBIR tool for characterizing various orbital lesions. Furthermore, we found evidence for a decrease in reading times when using only the CBIR tool, but an increase in reading time when using CBIR in conjunction with status quo tools.

Our measured diagnostic accuracy of 55.88% with status quo reference tools is comparable to other studies that assessed accuracy for orbital lesions^2,18^. However, our measured status quo accuracy is considerably higher than status quo measurements of most studies that analyzed the effect of CBIR on interstitial lung disease diagnostics in chest CT. There, the reported diagnostic accuracies range between 35%^7^ and 46.1%^5^, except for Pogarell et al.^8^ who reported 30% for novice and 60.7% for resident readers. The positive effect of the CBIR tool on diagnostic accuracy is comparable to the effects reported in Choe et al.^5^ (without CBIR 46.1%, with CBIR 60.9%), but more moderate than the ones reported in other studies^7,8^. In general, the measured diagnostic accuracy in our and other studies might underestimate the true diagnostic accuracy in the clinic, as only limited patient history and no laboratory data, nor reports from other sub-specialties were available to the participants.

The effect of CBIR on reading time is mixed in the literature. Haubold et al. find an increase in reading time by 22% (p<0.001) which moderates to 7% after readers become more familiar with the software^7^, whereas Röhrich et al. find a decrease by 31.3% (p<0.001)^6^. In our study we found a significant 29% decrease in reading times when using only the CBIR tool, and a significant 19% increase when SQ+CBIR tools were used for diagnosing eye and orbit mass lesions. Other studies did not analyze whether the CBIR tool was used in conjunction with other tools, thus the two opposing effects could be conflated. However, our study may have overestimated reading times with the CBIR tool, since participants only read four cases having the CBIR tool available, thus they only had limited time to get used to the software and reading times might be lower under routine conditions.

In other studies, participants were required to read 54^6^ or more cases in total^5,7,8^, which allows readers to become more familiar with the software but severely limits the total number of study participants that could be included to 8^5,6^ or less^7,8^. In our study, the low number of cases per participant allowed us to include 36 participants with considerable differences in experience and tenure, which better accounts for the heterogenous effects that AI assistance can have on radiologists^19^. In addition, this and other studies^5,6^ compared CBIR usage with status quo reference tools, whereas others compared CBIR assistance to no assistance at all^7,8^, which may lead to different interpretations of the impact of CBIR tools on outcome variables.

This study has two main limitations. While we included a diverse range of cases and participants, the small sample size still limits the generalizability of our findings. Further studies will expand to a larger and more geographically diverse participant and case pool, ideally involving participants from multiple medical centers, which would provide more robust data and would allow for more granular sub-group analyses. Another concern is the potential for the CBIR tool to negatively influence radiologists by retrieving confusing or irrelevant cases, which was not evaluated. Given that 5 of 36 participants and 9 of 48 cases had lower diagnostic accuracy with the CBIR tool available than without, it is crucial to assess if there exist underlying systematic factors, either radiologist-specific or case-specific, that may lead to this disparate impact. Prior AI research suggests that radiologists’ decisions can be influenced by AI errors and that this effect is more severe for inexperienced radiologists^20^. A similar effect could occur with CBIR if retrieved cases are misinterpreted as strong diagnostic evidence. In contrast, our results suggest that inexperienced radiologists gain the most and have the highest diagnostic accuracy across experience levels when having the CBIR tool available (**Tab. 3 d, Suppl. Fig 3 a**). In addition, we do not find a clear relationship between model retrieval performance scores for individual pathologies and diagnostic accuracy of study participants (**Suppl. Fig. 3 d**). Future work should examine how CBIR influences diagnostic reasoning and whether retrieval-based recommendations affect radiologists differently depending on experience levels and retrieval quality. Furthermore, mitigation strategies to reduce the risk of over-reliance of radiologists on CBIR outputs should be assessed. Potential approaches include integrating uncertainty or confidence scores alongside retrieved images to help users gauge retrieval reliability, providing guidelines on how to critically interpret CBIR results, and implementing feature importance heatmaps overlaid on query images to highlight key regions driving similarity scores^21^.

In conclusion, adopting CBIR in routine diagnostic workflows for eye and orbital mass lesions could have a substantial positive impact on radiological decision making and thus patient outcomes. However, more work is needed to assess the benefits of CBIR tools in other organ systems and imaging modalities. We plan to continue developing and refining the CBIR tool, expanding it to other organ systems and testing it in future studies.

## Data Availability

Measurements from the reader study are available from the corresponding author upon request.

## Acknowledgements

We thank our participants for their time and dedication. We are grateful for the support from Robert Röhle from the Charité Institute for Biometrics for giving advice on the statistical analysis. K.E.E., W.L., S.S. and J.L.R. received funding from the Digital Health Accelerator of the Berlin Institute of Health. K.E.E. received support from Stiftung Charité. J.L.R. received support from the IFI program of the German Academic Exchange Service (DAAD).

## Author Contributions

J.L.R. and K.E.E. conceived the study. W.L., A.L., B.W., A.S. and A.-S.I. retrieved the data. W.L. and S.-S.I. annotated the data. J.L.R and S.S. developed and deployed the software together with external service providers. W.L., B.W. and E.B.S conducted the experiments with the participants. J.L.R. did the statistical analysis. J.L.R. and K.E.E. wrote the draft manuscript. All authors revised the manuscript.

## Additional Information

The software used in this study was developed for research purposes but may be commercially licensed in the future, which may benefit authors J.L.R., W.L, S.S., S.S.I and K.E.E. None of the other authors declare competing interests.

