## Supplement for "Content-based image retrieval assists radiologists in diagnosing eye and orbital mass lesions in MRI"

**Supplementary Material**

| **Table 4: Reading time with/out CBIR** | | | | | | |
| --- | --- | --- | --- | --- | --- | --- |
| Characteristics | N | Only SQ | Only CBIR | P value | SQ+CBIR | P value |
| A) Difficulty |  |  |  |  |  |  |
| Easy | 105 | 202$\pm$113 | 158$\pm$78 | .46 | 260$\pm$173 | .44 |
| Hard | 138 | 357$\pm$206 | 271$\pm$198 | .002* | 462$\pm$212 | .03* |
| Really hard | 39 | 464$\pm$317 | 364$\pm$144 | .41 | 428$\pm$215 | .29 |
| B) Pathology Type |  |  |  |  |  |  |
| Infl. & Infect. | 102 | 338$\pm$201 | 226$\pm$112 | .005* | 389$\pm$242 | .10 |
| Benign | 54 | 363$\pm$240 | 335$\pm$304 | .32 | 476$\pm$278 | .83 |
| Malignant | 132 | 314$\pm$250 | 207$\pm$126 | <.001* | 365$\pm$163 | .045* |
| C) Medical Role |  |  |  |  |  |  |
| Resident | 128 | 417$\pm$278 | 276$\pm$232 | <.001* | 441$\pm$223 | .61 |
| Board-certified | 80 | 208$\pm$96 | 228$\pm$136 | .34 | 205$\pm$64 | .79 |
| Senior | 80 | 273$\pm$112 | 195$\pm$90 | .07 | 360$\pm$188 | .87 |
| D) Prior Experience |  |  |  |  |  |  |
| No exp. | 56 | 313$\pm$160 | 288$\pm$181 | .75 | 393$\pm$140 | .19 |
| Little exp. | 168 | 377$\pm$263 | 236$\pm$186 | .02* | 397$\pm$232 | .77 |
| Sufficient exp. | 64 | 204$\pm$113 | 194$\pm$122 | .80 | - |  |
| E) Tenure |  |  |  |  |  |  |
| 0-5 years | 128 | 417$\pm$278 | 276$\pm$232 | <.001* | 441$\pm$223 | .049* |
| 6-10 years | 80 | 237$\pm$104 | 210$\pm$126 | .006* | 408$\pm$179 | .89 |
| 11-15 years | 32 | 187$\pm$89 | 250$\pm$133 | .002* | - |  |
| >15 years | 48 | 286$\pm$114 | 188$\pm$74 | .16 | 198$\pm$57 | .25 |
| F) Overall |  |  |  |  |  |  |
| All | 288 | 334$\pm$230 | 236$\pm$172 | <.001* | 396$\pm$215 | <.001* |

Note. — Statistics of reading time in seconds are shown for measurements where only status quo reference tools were used (Only SQ), where only the CBIR tool was used (Only CBIR) and where both were used (SQ+CBIR). P values indicate significant differences to reference level ‘Only status quo’ and were calculated using linear mixed effects models with individual readers and patients as random effects.

**Content-based image retrieval assists radiologists in diagnosing rare eye and orbital mass lesions in MRI**

J. Lorenz Rumberger^1,2,3,†^, Winna Lim^4,†^, Benjamin Wildfeuer^4^, Elisa B. Sodemann^4^, Augustin Lecler^5,6^, Simon Stemplinger, Ahi Sema-Issever^4^, Ali R. Sepahdari^7,8^, Sönke Langner^9^, Dagmar Kainmueller^1,3,10^, Bernd Hamm^4^, Katharina Erb-Eigner^4,*^

^1^Max-Delbrück Center for Molecular Medicine in the Helmholtz Association, Berlin, Germany; ^2^Humboldt University Berlin, Faculty of Mathematics and Natural Sciences, Berlin, Germany; ^3^Helmholtz Imaging; ^4^Charité-Universitätsmedizin Berlin, Corporate Member of Freie Universität Berlin and Humboldt Universität zu Berlin, Berlin, Germany; ^5^Hôpital Fondation Adolphe de Rothschild, Paris, France; ^6^Paris Cité University, Paris, France; ^7^Diagnostic Neuroradiology, Department of Radiology, Scripps Clinic Medical Group, La Jolla, California, USA; ^8^David Geffen School of Medicine, University of California Los Angeles, Los Angeles, California, USA; ^9^Greifswald University Medicine, Greifswald, Germany; ^10^Potsdam University, Digital Engineering Faculty, Potsdam, Germany

| Characteristics | Only CBIR | P value | SQ+CBIR | P value |
| --- | --- | --- | --- | --- |
| **a)** Difficulty |  |  |  |  |
| Easy | 5.69 (1.25 – 25.96) | 0.02* | 3.89 (0.00 – Inf) | 0.99 |
| Hard | 1.36 (0.59 – 3.11) | 0.47 | 2.27 (0.60 – 8.61) | 0.23 |
| Really hard | 1.15 (0.26 – 5.08) | 0.86 | 7.19 (0.58 – 89.71) | 0.13 |
| **b)** Pathology Type |  |  |  |  |
| Infl. & Infect. | 2.97 (0.97 – 9.02) | 0.055 | 4.38 (0.73 – 26.13) | 0.11 |
| Benign | 0.94 (0.24 – 3.64) | 0.93 | 6.87 (0.59 – 79.51) | 0.12 |
| Malignant | 1.88 (0.69 – 5.13) | 0.22 | 2.90 (0.52 – 16.18) | 0.23 |
| **c)** Medical Role |  |  |  |  |
| Resident | 2.46 (0.86 – 7.04) | 0.09 | 2.87 (0.78 – 10.58) | 0.11 |
| Board-certified | 0.73 (0.21 – 2.55) | 0.62 | 1.97 (0.14 – 28.14) | 0.62 |
| Senior | 4.69 (1.40 – 16.33) | 0.01* | Inf (0.00 – Inf) | 0.99 |
| **d)** Prior Experience |  |  |  |  |
| No exp. | 3.30 (0.70 – 12.03) | 0.10 | Inf (0.00 – Inf) | 0.99 |
| Little exp. | 1.82 (0.80 – 4.17) | 0.15 | 3.16 (0.96 – 10.37) | 0.058 |
| Sufficient exp. | 1.21 (0.31 – 4.77) | 0.79 |  |  |
| **e)** Tenure |  |  |  |  |
| 0-5 years | 2.38 (0.85 – 6.66) | 0.10 | 2.89 (0.79 – 10.55) | 0.11 |
| 6-10 years | 2.43 (0.73 – 8.05) | 0.15 | Inf (0.00 – Inf) | 0.90 |
| 11-15 years | 0.63 (0.09 – 4.25) | 0.64 |  |  |
| >15 years | 3.42 (0.61 – 19.31) | 0.16 | 2.80 (0.20 – 38.99) | 0.44 |
| **f)** Overall |  |  |  |  |
| All | 2.07 (1.08 – 3.95) | 0.03* | 3.65 (1.21 – 3.95) | 0.02* |

**Suppl. Tab. 1 | Odds Ratios for Diagnostic Accuracy with CBIR.** Odds ratios of diagnostic accuracies for measurements where only the CBIR tool was used (Only CBIR) and when it was used together with status quo tools (SQ+CBIR), relative to reference level only status quo (Only SQ). Odds ratios and P values were calculated by using logistic mixed effects models with individual readers and patients as random effects. 95%- Wald confidence interval reported in parenthesis.

| Characteristics | Only CBIR | P value | SQ+CBIR | P value |
| --- | --- | --- | --- | --- |
| **a)** Difficulty |  |  |  |  |
| Easy | 0.84 (0.68 – 1.05) | 0.14 | 1.58 (1.12 – 2.23) | 0.01* |
| Hard | 0.76 (0.64 – 0.89) | 0.002* | 1.33 (1.04 – 1.71) | 0.03* |
| Really hard | 0.88 (0.65 – 1.19) | 0.41 | 1.02 (0.66 – 1.56) | 0.94 |
| **b)** Pathology Type |  |  |  |  |
| Infl. & Infect. | 0.73 (0.58 – 0.90) | 0.005* | 1.33 (0.95 – 1.85) | 0.10 |
| Benign | 0.88 (0.64 – 1.18) | 0.40 | 1.25 (0.80 – 1.97) | 0.34 |
| Malignant | 0.71 (0.58 – 0.86) | <0.001* | 1.37 (1.01 – 1.85) | 0.045* |
| **c)** Medical Role |  |  |  |  |
| Resident | 0.58 (0.47 – 0.70) | <0.001* | 1.28 (1.00 – 1.61) | 0.046* |
| Board-certified | 1.15 (0.87 – 1.53) | 0.34 | 1.08 (0.61 – 1.94) | 0.79 |
| Senior | 0.80 (0.63 – 1.02) | 0.08 | 1.14 (0.72 – 1.83) | 0.58 |
| **d)** Prior Experience |  |  |  |  |
| No exp. | 0.95 (0.71 – 1.27) | 0.75 | 1.36 (0.87 – 2.11) | 0.19 |
| Little exp. | 0.64 (0.53 – 0.76) | <0.001* | 1.26 (0.99 – 1.58) | 0.054 |
| Sufficient exp. | 0.90 (0.67 – 1.21) | 0.50 |  |  |
| **e)** Tenure |  |  |  |  |
| 0-5 years | 0.58 (0.48 – 0.71) | <0.001* | 1.27 (1.00 – 1.61) | 0.049* |
| 6-10 years | 0.93 (0.72 – 1.19) | 0.58 | 1.33 (0.80 – 2.25) | 0.29 |
| 11-15 years | 1.24 (0.82 – 1.90) | 0.32 |  |  |
| >15 years | 0.78 (0.55 – 1.10) | 0.17 | 0.91(0.54 – 1.53) | 0.73 |
| **f)** Overall |  |  |  |  |
| All | 0.75 (0.66 – 0.86) | <0.001* | 1.32 (1.07 – 1.61) | <0.001* |

**Suppl. Tab. 2 | Adjusted impact of CBIR usage on Reading time**. Exponential of the regression coefficients of log(reading time) in seconds are shown for measurements where only the CBIR tool was used (Only CBIR) and where it was used together with status quo tools (SQ+CBIR). P values indicate significant differences to reference level ‘Only status quo’ and were calculated using linear mixed effects models with individual readers and patients as random effects. 95%- Wald confidence interval reported in parenthesis.

**S1: Definition of Clinically Significant vs. Insignificant Errors**

In our study, diagnostic errors were categorized as either clinically significant or clinically insignificant based on their potential impact on patient management. This distinction was essential for assessing diagnostic accuracy and understanding the practical implications of classification errors.

Clinically significant errors were defined as misclassifications that could lead to substantial differences in treatment decisions or patient outcomes. These errors typically involved cases where two entities had distinct imaging characteristics and required different clinical management. One example of a clinically significant error is the misclassification of rhabdomyosarcoma as lymphoma. While both are malignant tumors, rhabdomyosarcomas and lymphomas exhibit different imaging features and necessitate distinct treatment strategies. A misdiagnosis in such cases could result in inappropriate management, making it a significant error in the context of patient care.

In contrast, clinically insignificant errors were cases where different but closely related terms were used without affecting the overall diagnostic interpretation or treatment decision. These errors occurred within broad diagnostic categories where multiple descriptors were considered acceptable. An example of a clinically insignificant error is related to the classification of non-specific orbital inflammatory disease. Many inflammatory orbital diseases are grouped under this broad category. If a reader used more specific but related terms, such as cellulitis, scleritis, or dacryoadenitis, these were still considered correct since they indicated an inflammatory origin and did not alter the clinical approach.

**S2: Data pre-processing steps**

The image pre-processing pipeline for CBIR development and fine-tuning incorporated normalization, data augmentation, and sampling strategies to improve model robustness and generalization. First, images underwent volume-wide percentile normalization, ensuring intensity values were standardized. To focus on regions relevant for diagnosis, only images containing the pathology (which was before annotated with a bounding box) were retained, while others were excluded from training.

To address class imbalances, importance sampling was applied during training, where rare classes were oversampled and abundant classes were undersampled, resulting in a more uniform class distribution. A range of geometric transformations was introduced to enhance data diversity, including random affine transformations with rotation, translation, scaling, as well as horizontal and vertical flipping. Additionally, elastic deformations were used to simulate anatomical variability. Intensity-based augmentations included random contrast and brightness adjustments, motion blur to replicate variations in imaging conditions, and Rician noise to model MRI-specific distortions.

Furthermore, a random view transformation was applied, generating multiple views of each image while ensuring that the pathology remained visible. This helped the model focus on diagnostically relevant structures while preventing it from learning biases introduced by inconsistent imaging perspectives.

**S3: Pre-training datasets**

For training the DinoV2 model, we curated a diverse collection of radiological images, focusing on MRI and CT scans while filtering out color images to maintain consistency. Even though some datasets included text captions, we exclusively used image data for DinoV2 pre-training. The datasets utilized include:

- MPx-Single: A subset of the MedPix dataset, which contains radiological images with annotations on scan-level such as image modality, shooting plane, and textual descriptions.
- VQA-RAD: A dataset of visual question-answering (VQA) pairs generated from radiology reports, covering various imaging modalities, including X-rays, CTs, and MRIs.
- RP3D: The Radiopaedia 3D dataset, which includes a diverse collection of annotated 3D radiological images, supporting captioning and question-answering tasks.
- PMC-OA: The PubMed Central Open Access (PMC-OA) dataset, comprising radiological images extracted from PMC Open Access articles.

**S4: Model Performance**

Despite the relatively small fine-tuning dataset, the CBIR model performs well, especially on the validation set. There, precision@k for k=5, 10, 20 ranges from 0.24 to 0.26, and top-k accuracy reaches 0.55 at k=10. Performance is lower on the external test set (precision@k around 0.13, top-10 accuracy up to 0.48). However, the model still retrieves relevant cases in many instances**.** The performance difference between the validation and external test datasets is present but less pronounced when considering **top-k accuracy**, which better reflects retrieval quality given the pathology-based grouping of search results in the interface (c.f. **Fig. 3 b**). Note that the reader study dataset was sourced largely from Charité, like the validation dataset, meaning that the validation performance (**Suppl. Fig 1 c-d**) better reflects the CBIR effectiveness within the reader study setting than the external test set performance (**Suppl. Fig 1 a-b**).

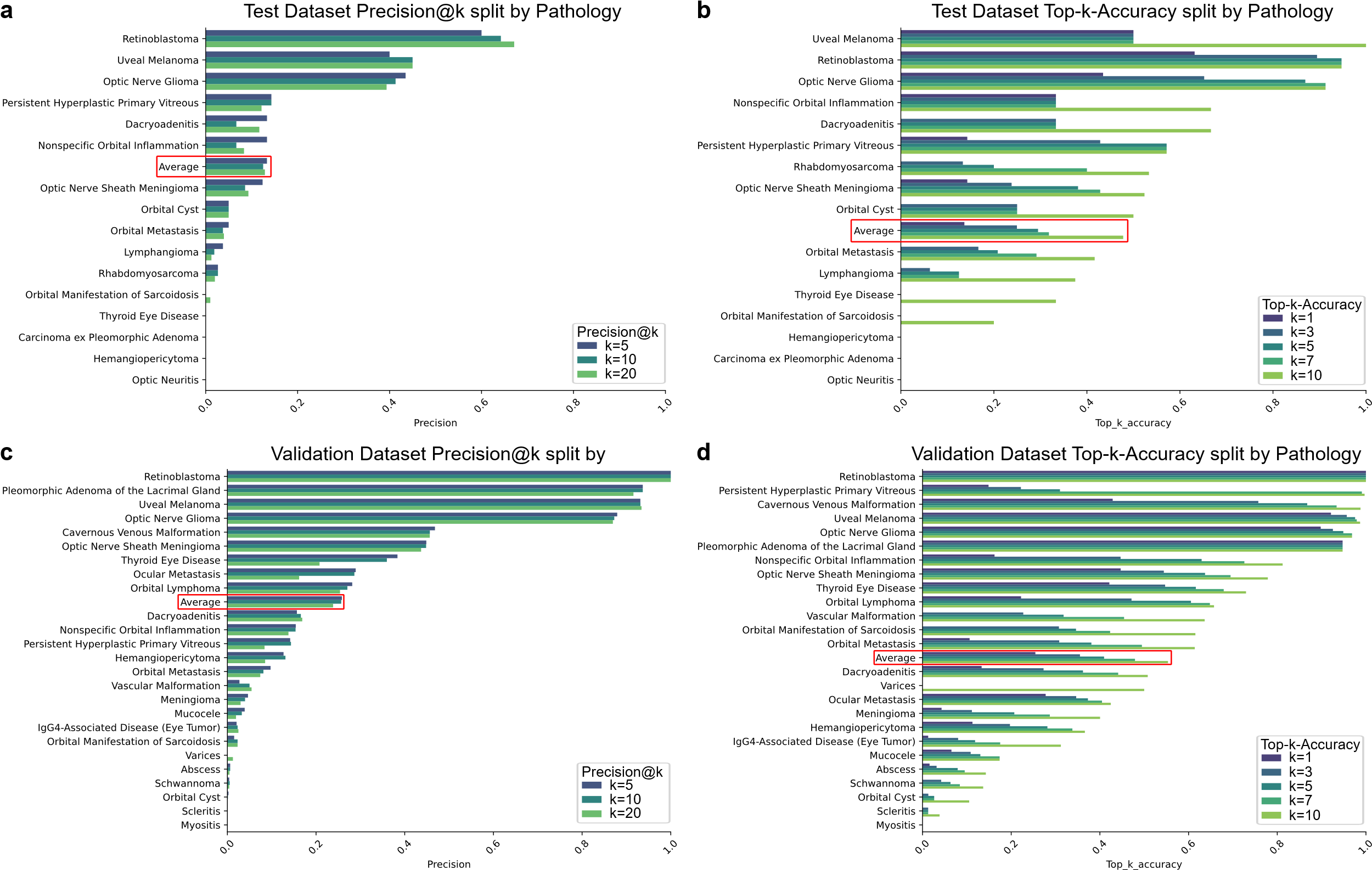
 **Suppl. Fig. 1 | Model Performance .** Precision@k metric split by pathology for the test dataset (**a**) and for the validation dataset (**c**). Top-k-Accuracy metric split by pathology for the test dataset (**b**) and for the validation dataset (**d**), averages over the individual pathologies highlighted in red.

**S5: Discussion of Cases where accuracy dropped to 0% with CBIR**

Three cases showed a significant drop in diagnostic accuracy when CBIR was available (**Fig. 4 a**), falling from 66.66% (without CBIR) to 0% (with CBIR). These cases had the ground truth diagnoses Arteriovenous Malformation (AVM), Lymphoma, and Preseptal Cellulitis.

This decline suggests CBIR may have retrieved visually similar but clinically irrelevant cases, which may mislead the study participants. The variable vascular patterns of AVMs, the diverse imaging presentations of Lymphomas, and the similarity to other inflammatory conditions of preseptal cellulitis could have contributed to incorrect case matches. Additionally, over-reliance on CBIR may have influenced radiologists to trust retrieved cases over their own expertise. A key limitation is the lack of additional context awareness of the CBIR system, as it retrieves images based on visual features alone without considering patient history or additional information. It may be beneficial to consider these other sources of information to further improve the relevance of the search results.

**S6: Cases where no reference tool was used**

| **Characteristics** | **Diagnostic Accuracy** |
| --- | --- |
| **a \|** **Difficulty** |  |
| Easy | 87.88 (29/33) |
| Hard | 68.42 (13/19) |
| Really hard | 0.00 (0/1) |
| **b \|** **Pathology Type** |  |
| Infl. & Infect. | 73.68 (14/19) |
| Benign | 57.14 (4/7) |
| Malignant | 89.29 (25/28) |
| **c \|** **Medical Role** |  |
| Resident | 86.67 (13/15) |
| Board-certified | 68.18 (15/22) |
| Senior | 88.24 (15/17) |
| **d \|** **Prior Experience** | |
| No exp. | 100.00 (3/3) |
| Little exp. | 76.67 (23/30) |
| Sufficient exp. | 80.95 (17/21) |
| **e \|** **Tenure** |  |
| 0-5 years | 86.67 (13/15) |
| 6-10 years | 66.67 (12/18) |
| 11-15 years | 81.82 (9/11) |
| >15 years | 90.00 (9/10) |
| **f \|** **Overall** |  |
| All | 79.63 (43/54) |

**Suppl. Tab. 3 | Diagnostic accuracy for cases where no references were used.**

Unless otherwise stated, data is presented as percentages relative to the total number of measurements. Fractions of total number of measurements in parenthesis.

We analyzed the performance of participants who did not use reference tools (**Suppl. Tab. 3**). Overall, 79.63% of diagnoses were correct, with high accuracy in easy (87.88%) and intermediate accuracy in hard cases (68.42%). Accuracy was highest for malignant (89.29%), followed by inflammatory/infectious (73.68%) and benign (57.14%) pathologies. Results across medical roles, tenure levels and prior experience are mixed with no clear trend that higher ranked roles or additional tenure clearly increases diagnostic accuracy when no reference tools were used.

However, there is a strong selection effect -participants only refrained from using a reference tool when they immediately recognized the diagnosis with high confidence. This means their performance in these cases cannot be directly compared to instances where reference tools were used, as those cases inherently included greater diagnostic uncertainty. Thus, while high accuracy was observed in this subgroup, it does not imply that not using a reference tool led to better performance, but rather that these were the cases where external support was deemed unnecessary by the participants.

**S7: Power analysis and Case Number Calculations**

The power analysis aimed to determine the minimum effect size detectable given the study design, participant count, and statistical power. We followed the approach used in Röhrich et al., who studied diagnostic accuracy and reading time improvements in a similar setting with 8 participants reading 54 cases each, resulting in 432 total measurements. Their findings included a 31.3% decrease in reading time (p<0.001, Cohen’s d > 3) and a non-significant improvement in diagnostic accuracy (34.7% to 42.2%, p=0.083). For our study, we targeted 36 participants reading 6-8 cases each, leading to 160-288 total measurements, depending on feasibility. The number of cases per participant was informed by 4 dry runs to ensure that the total study duration remained below 60 minutes per participant. Given the variability of cases and expected smaller effect sizes (approximately Cohen’s d = 1.0), we opted for 36 participants and 48 cases, which allowed us to detect effects as small as Cohen’s d = 0.6 with 80% statistical power (**Suppl. Fig. 2**). To account for potential variance between cases and participants, we implemented 6 repeated measurements per case, and 8 measurements per participant to ensure robust estimation of random effects.

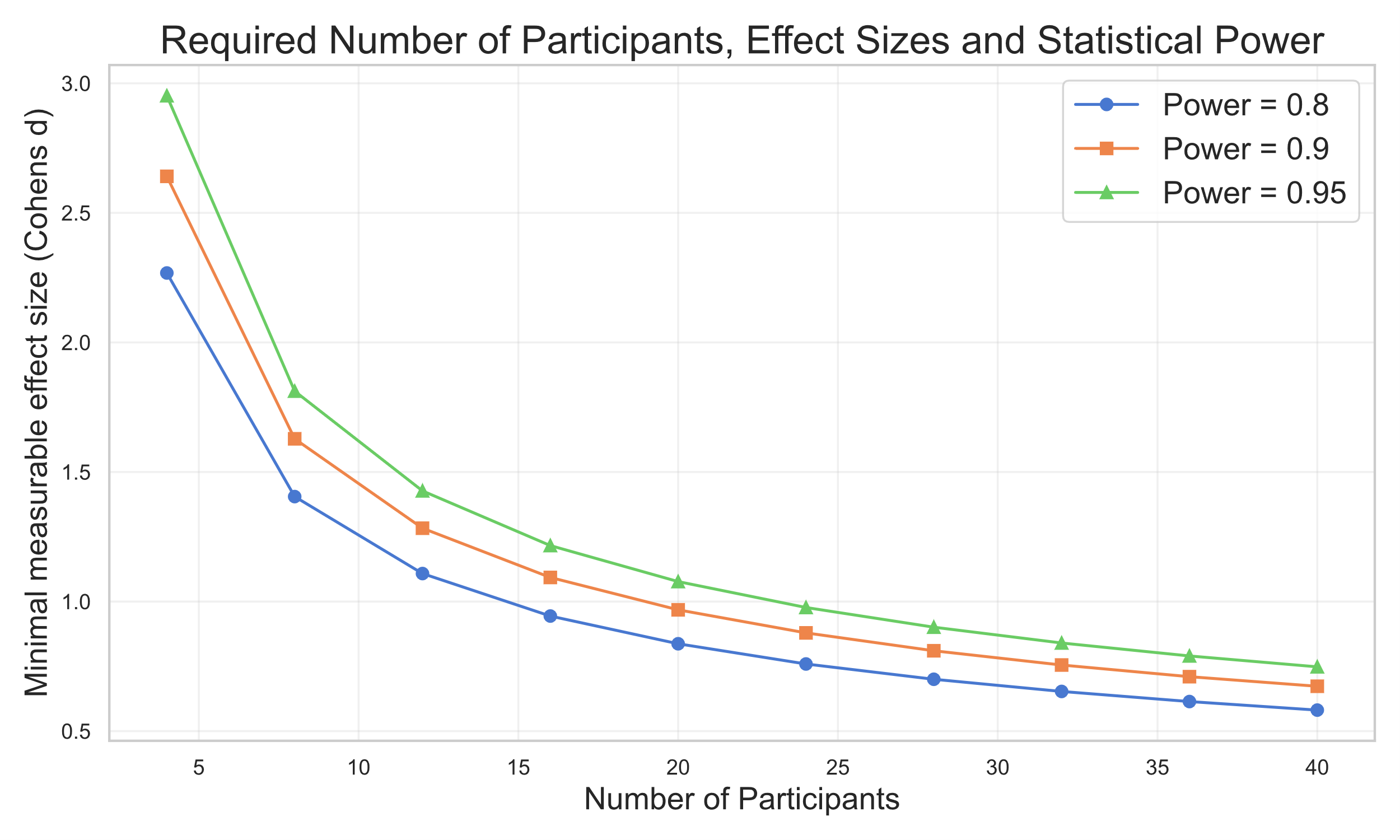
**Suppl. Fig. 2 | Power analysis.** Minimal measurable effect sizes with Mixed Effects Models for variable numbers of participants.

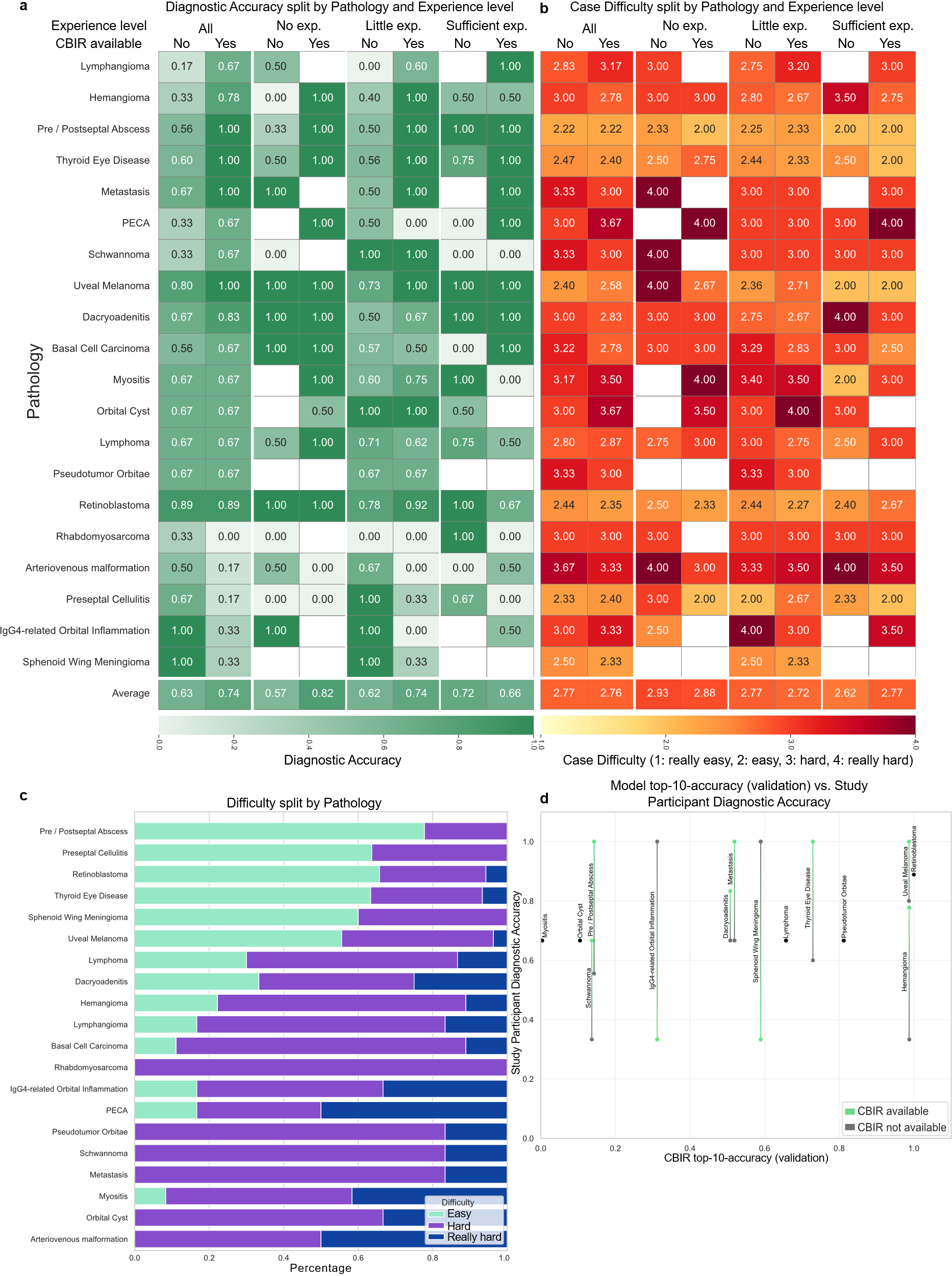
**Suppl. Fig. 3 | Accuracy and Difficulty Ratings split by Pathology. a-b**, heatmaps showing average diagnostic accuracy (**a**) and difficulty scores (**b**) for each pathology across experience levels. **c**, stacked bar chart displays the overall distribution of difficulty ratings. **d**, shows the top-10-accuracy of the CBIR tool on the validation dataset (x-axis) and the diagnostic accuracy of study participants (y-axis) for cases when the CBIR was available (green) or not (gray) and the difference between the two as a bar, when there was no difference, dots are colored black.
